# Large Language Model Extraction and Inference of Physician Global Assessment from Clinical Notes in Juvenile Idiopathic Arthritis

**DOI:** 10.64898/2026.11.22.26368678

**Authors:** Mei-Sing Ong, Jiarui Yao, Seungwon Lee, Guergana Savova, Marc D. Natter

## Abstract

**Background:** The physician global assessment (PhGA) is a core outcome measure in juvenile idiopathic arthritis (JIA) but is incompletely captured, even in clinical registries. We developed a large language model (LLM) approach that extracts explicitly documented PhGA from clinical narratives and infers a score where none is recorded. To our knowledge, this is the first study to address both tasks.

**Methods:** We linked charts from 306 children with JIA at one center to the Childhood Arthritis and Rheumatology Research Alliance (CARRA) Registry (2016–2024). A structured prompt was developed on 100 randomly sampled charts, then evaluated on 400 charts from patients not represented in the development set. Extraction used GPT-5.2 within a HIPAA-compliant Azure environment. Registry-recorded PhGA served as the reference standard, with discordances adjudicated by a pediatric rheumatologist blinded to both sources. Agreement was assessed by intraclass correlation coefficient (ICC(2,1)), exact match, and mean absolute error (MAE).

**Results:** Against registry values, ICC(2,1) was 0.87 (95% CI 0.85–0.89; n=400), with 70.5% exact agreement and MAE 0.363. Agreement was near-perfect for explicit extractions (n=190; ICC 0.99; 95% CI 0.98-0.99) and lower for inferred scores (n=210; ICC 0.72; 95% CI 0.64-0.78). Large discordances (≥2 points) occurred in 7.3% of cases. Against the adjudicated standard, overall ICC(2,1) was 0.96 (95% CI 0.95–0.97), with 77.5% exact agreement and MAE 0.200. All 14 explicit discordances reflected mismatch between the registry and the clinical note, rather than extraction errors (adjudicated ICC 1.00). For inferred scores, ICC rose to 0.90 (95% CI 0.87-0.92). Residual disagreement was concentrated at the inactive/minimally active boundary.

**Conclusion:** An LLM can reliably extract PhGA from clinical notes with high accuracy to support downstream clinical research, offering a scalable approach to mitigating missingness in patient registries.

**Funding Support:** Patient-Centered Outcomes Research Institute (PCORI) Award (ME-2022C2-25573- IC). The study’s funder played no role in the study design, interpretation of findings, writing of the report, or the decision to submit this manuscript for publication.

## INTRODUCTION

The physician’s global assessment of disease activity (PhGA) is a physician assigned score that reflects a clinician’s overall assessment of disease activity, with values ranging from 0 for no activity, to 10 for maximum activity. It is a core outcome measure in juvenile idiopathic arthritis (JIA), serving as a component of the American College of Rheumatology JIA response criteria for defining clinically inactive disease.^1^ Despite its importance, PhGA, which should be assigned by the practitioner at or near the conclusion of a clinic visit, is not consistently recorded at every encounter and is incompletely captured in clinical registries, limiting the completeness of disease activity measures and the cohorts available for longitudinal analysis. In the Childhood Arthritis and Rheumatology Research Alliance (CARRA) Registry – the largest clinical registry for childhood rheumatic diseases in North America, PhGA was missing from 6% of visits at baseline, rising to 82% at the 36-month follow-up.^2^ Nonetheless, PhGA scores are often documented in clinical notes, either explicitly or as reflected in the treating rheumatologist’s description of the physical examination and overall clinical impression.

With the advent of large language models (LLMs), there is a new opportunity not only to extract explicitly documented values from clinical text, but also to infer values that are not directly stated in the clinical record. The latter requires integrating contextual information captured in the clinical narrative, a capability that is difficult to achieve with traditional natural language processing approaches. We hypothesized that frontier LLMs could be effective in retrospectively inferring PhGA scores from clinical narratives, processing the treating rheumatologist’s history, physician examination, and impression/plan. Here, we developed an LLM-based approach to extract PhGA from clinical narratives, extracting explicitly documented PhGA values and inferring a score from the narrative where none is recorded. To our knowledge this is the first study to address both extraction and inference of PhGA, offering a practical route to reducing missingness in registry-based disease activity monitoring.

## METHODS

### Study cohort and dataset

The study cohort comprised 306 children with JIA seen at Boston Children’s Hospital (BCH) who were also enrolled in the Childhood Arthritis and Rheumatology Research Alliance (CARRA) Registry (years 2016-2024) (Table 1). The Registry captures gold standard PhGA data reported by the treating pediatric rheumatologists. We extracted clinical notes from the electronic health record (EHR) at BCH and linked each note to its corresponding CARRA Registry visit. A random sample of 100 charts were selected for prompt development before evaluation on a 400-chart test set drawn from patients not represented in the development set to prevent data leakage. Subjects contributed a mean of 1.6 charts each (median 1, range 1-6).

**Table 1.** Study participants (n=304)

| **Attribute** |  |
|---|---|
| Age in years, median (IQR) | 12 (7 – 15) |
| Gender, n (%) |  |
| Female | 199 (65.5) |
| Male | 98 (32.2) |
| Not reported | 7 (2.3) |
| JIA category, n (%) |  |
| Enthesitis-related arthritis | 31 (10.2) |
| Oligoarthritis | 84 (27.6) |
| Polyarthritis (Rheumatoid factor negative) | 75 (24.7) |
| Polyarthritis (Rheumatoid factor positive) | 21 (6.9) |
| Psoriatic arthritis | 56 (18.4) |
| Systemic arthritis | 29 (9.5) |
| Undifferentiated arthritis | 8 (2.6) |

This study was approved by the Boston Children’s Hospital Institutional Review Board (IRB- P00030533).

### Prompt development

A structured prompt template was developed to extract PhGA from clinical narratives. The prompt instructs the LLM to first identify a physician-documented numeric PhGA value when present in the note (i.e. “explicit” extraction), and if no explicit value can be detected, to derive PhGA from documented examination findings, disease activity narrative, and treatment context (i.e. “inferred” extraction).

Inference of PhGA was guided by scoring rules based on joint count, site-specific findings (e.g., temporomandibular joint, dactylitis), and presenting symptoms. Treatment direction (e.g. escalation of therapy) and laboratory findings were applied as secondary modifiers. Output was returned in structured format including the extracted PhGA value, the extraction type (i.e. explicit vs inferred), a confidence rating (high, medium or low), supporting text (verbatim text quoted from the note as evidence) and rationale (a brief explanation of how the score was derived).

The template was iteratively refined across multiple versions based on chart-level error analysis in the development set, with the final version locked prior to evaluation on the test set. Iterations addressed specific error patterns identified through manual chart review. These included explicit handling of near- remission presentations with minor residual exam or symptom findings to avoid systematic under-scoring of near remission cases; recognition of extra-articular features in the absence of active joint (e.g. uveitis, psoriasis); recognition of additional PhGA label conventions for explicit value detection; and consideration of treatment direction as a modifier.

PhGA extraction and inference were performed using GPT-5.2 (OpenAI), the most advanced frontier model available within BCH’s HIPAA-compliant Microsoft Azure Service environment at the time of the study. Processing was conducted within this institutionally governed infrastructure, ensuring that clinical note content did not leave institutional control. The reference standard was the CARRA registry- documented PhGA, supplemented by blinded adjudication by a pediatric rheumatologist (MDN).

Accordingly, all discordances of magnitude ≥2 between LLM-extracted and registry-documented values were reviewed by a pediatric rheumatologist masked to both registry-documented and LLM-extracted PhGA. All explicit-extraction discordances were manually reviewed regardless of magnitude. Analyses were stratified by extraction modes (explicit versus inferred) and by the LLM’s self-reported confidence level. Primary agreement statistics were intraclass correlation coefficient (ICC(2,1); two-way random- effects, single-measure, absolute agreement), mean absolute error (MAE), mean signed difference (i.e. measure of directional bias), and percentage exact/near match to the reference standard. Provider-level variability was examined as an exploratory sub-analysis.

### Self-consistency evaluation

It is well-established that LLM outputs are not fully deterministic.^3^ The same prompt and input can result in different output across independent runs.^4^ Variation in model output has important implications for the repeatability of the extraction approach. We therefore conducted a separate analysis to quantify variability in chart-level scores across 3 independent runs on the test set, using the locked prompt with no modifications between runs and with all model parameters held constant. Agreement among runs was characterized by ICC(2,1) across runs, pairwise ICC(2,1) between each pair, the proportion of charts receiving an identical score across all runs, and the proportion with any disagreement in extraction type.

For each chart, we further computed the standard deviation of the extracted PhGA value across runs as a chart-level instability metric. This measure is independent of accuracy: a chart may be stably incorrect (low standard deviation, high error) or unstably correct. We conducted 2 exploratory comparisons using this metric: (a) whether chart-level instability is associated with LLM-reported extraction confidence; and (b) whether instability is associated with magnitude of disagreement from the reference standard.

### Multi-run aggregation

Given the inherent stochasticity of LLM outputs, aggregation across runs has the potential to reduce error where variation is stochastic.^5^ The median across runs was used as the aggregation method instead of the mean, since PhGA is recorded on a discrete 0.5-point grid and thus arithmetic averaging of discordant runs produces values that cannot exactly match any reference value. The median returns an observed value, and with three runs is unaffected by a single aberrant run.

### Assessment of hallucination

Accuracy of the extracted PhGA value does not establish that the LLM’s stated reasoning is faithful to the source note. An extraction may report a numerically correct score while citing a clinical finding that does not appear in the note, or referencing a scoring rule that does not exist in the prompt. We therefore conducted a separate assessment of extraction faithfulness. Accordingly, we assessed three potential categories of hallucination: (1) citation fabrication: one or more snippets in supporting text do not appear in the source note; (2) schema fabrication: rationale cites a rule that does not exist in the prompt; (3) rationale fabrication: rationale asserts a clinical finding that is not documented in the note, or that contradicts the note. An extraction was classified as containing a hallucination if it failed any of the three categories.

For citation fabrication, each snippet in supporting text was compared against the source note following Unicode normalization, harmonization of quotation-mark and dash variants, whitespace collapsing, and case-folding. A snippet was classified as grounded if it appeared as an exact normalized substring of the note. All flagged extractions underwent blinded manual review to determine whether they represented true hallucinations. In addition, an independent stratified random sample of 50 extractions (20 explicit, 30 inferred) underwent blinded manual review to provide an estimate for schema and clinical fabrications, and as an additional check on citation fabrication. The hallucination rate was reported, with 95% confidence intervals; where no events were observed, the upper bound of the 95% confidence interval is reported using the rule of three (3/n).^6^

## RESULTS

In the held-out test set, agreement between LLM-extracted and registry-documented PhGA was ICC(2,1) = 0.87 (95% CI 0.85–0.89). Extracted values matched the registry exactly in 70.5% of cases, with 81.8% falling within 0.5 points and 91.2% within 1.0 point (MAE 0.363). The mean signed difference was ™0.012, indicating no net directional bias overall. Figure 1 shows the distribution of the difference between LLM-extracted and registry-documented PhGA scores (LLM minus registry). Figure 2 shows the mean (SD) of this difference across each registry-documented PhGA score. Corresponding development set results, which informed prompt iteration are summarized in Table 2.

**Table 2.** Agreement between LLM-extracted/inferred and registry-documented PhGA, using the CARRA registry values as the reference standard.

|  | **All** | **Explicit** | **Inferred** |
|---|---|---|---|
| **Development set** |  |  |  |
| N | 100 | 47 | 53 |
| ICC(2,1) (95% CI) | 0.90 (0.85 – 0.93) | 0.91 (0.85 – 0.95) | 0.85 (0.75 – 0.91) |
| % Matching gold standard |  |  |  |
| Exact match | 60.0 | 83.0 | 39.6 |
| Within 0.5 | 74.0 | 83.0 | 66.0 |
| Within 1.0 | 92.0 | 93.6 | 90.6 |
| MAE | 0.425 | 0.287 | 0.547 |
| Mean signed diff (bias) | -0.135 | -0.245 | -0.038 |
| **Test set** |  |  |  |
| N | 400 | 190 | 210 |
| ICC(2,1) (95% CI) | 0.87 (0.85 – 0.89) | 0.99 (0.98 – 0.99) | 0.72 (0.64 – 0.78) |
| % Matching gold standard |  |  |  |
| Exact match | 70.5 | 92.6 | 50.5 |
| Within 0.5 | 81.8 | 96.3 | 68.6 |
| Within 1.0 | 91.2 | 98.4 | 84.8 |
| MAE | 0.363 | 0.068 | 0.629 |
| Mean signed diff (bias) | -0.012 | 0.016 | -0.038 |

**Figure 1.**
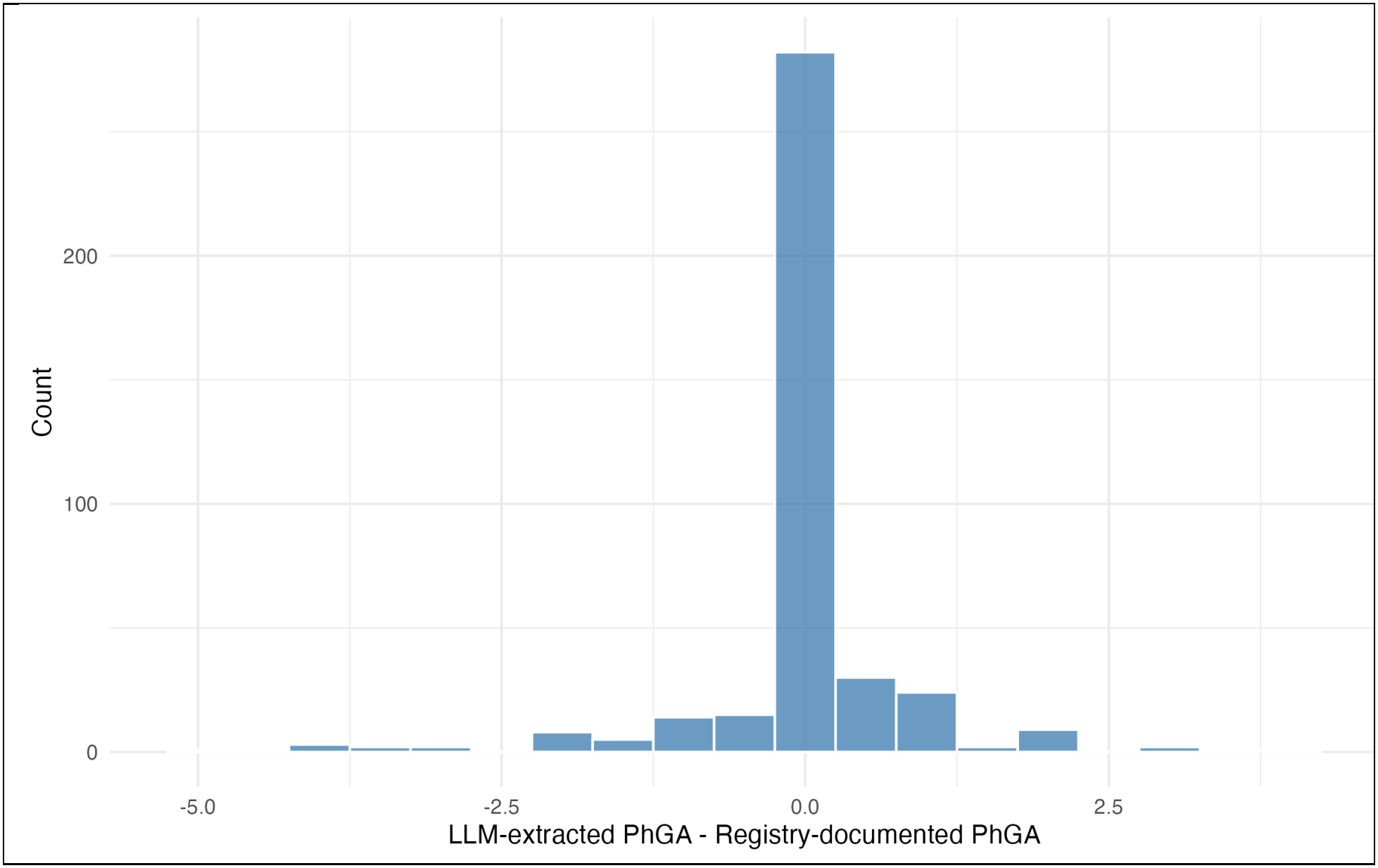
Distribution of the difference between LLM-extracted and registry-documented PhGA scores.

**Figure 2.**
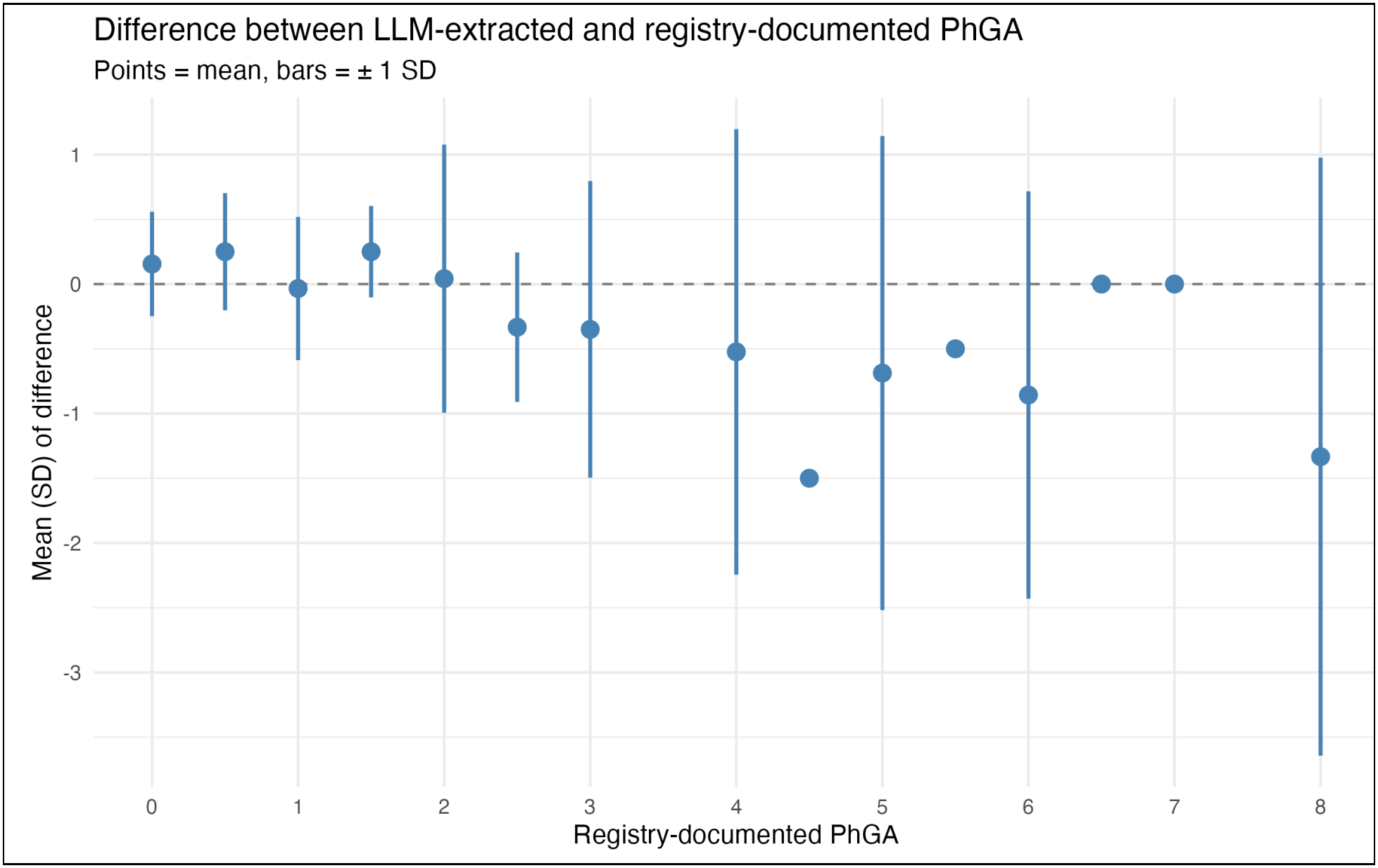
Difference between LLM-extracted/inferred and registry-documented PhGA scores (LLM minus registry) by registry-documented PhGA score. Points represent the mean and bars ± 1 standard deviation (SD). The dashed line indicates perfect agreement.

Large discordances (|difference| ≥ 2) were uncommon (5% [n=5] of development set and 7.2% [n=29] of test set comparisons) and were concentrated in inferred cases with medium LLM-reported confidence. In the test set, 41 cases were adjudicated: all 14 explicit discordances (with |difference| ≥ 2) and 27 inferred discordances with |difference| ≥ 2. Applying adjudicated values, ICC(2,1) rose to 0.96 (95% CI 0.95– 0.97), with exact agreement rising to 77.5% and MAE falling to 0.200 (Table 3).

**Table 3.** Agreement between LLM-extracted and registry-documented PhGA, using the CARRA registry values supplemented with blinded physician adjudication as the reference standard. All discordances with |LLM ™ registry| ≥ 2, and all explicit-extraction discordances of any magnitude, were adjudicated by a pediatric rheumatologist masked to value source. The adjudicated value replaced the registry value in the reference standard. “Number of corrections” indicates adjudicated cases assigned the LLM-extracted value.

|  | **All** | **Explicit** | **Inferred** |
|---|---|---|---|
| **Development set** |  |  |  |
| N | 100 | 47 | 53 |
| Number of corrections | 11 | 8 | 3 |
| ICC(2,1) (95% CI) | 0.94 (0.92 – 0.96) | 1.00 (1.00 – 1.00) | 0.86 (0.76 – 0.91) |
| % Matching gold standard |  |  |  |
| Exact match | 69.0 | 100 | 41.5 |
| Within 0.5 | 83.0 | 100 | 67.9 |
| Within 1.0 | 95.0 | 100 | 90.6 |
| MAE | 0.280 | 0 | 0.528 |
| Mean signed diff (bias) | -0.040 | 0 | -0.075 |
| **Test set** |  |  |  |
| N | 400 | 190 | 210 |
| Number of corrections | 41 | 14 | 27 |
| ICC(2,1) (95% CI) | 0.96 (0.95 – 0.97) | 1.00 (1.00 – 1.00) | 0.90 (0.87 – 0.92) |
| % Matching gold standard |  |  |  |
| Exact match | 77.5 | 100 | 57.1 |
| Within 0.5 | 87.8 | 100 | 76.7 |
| Within 1.0 | 97.0 | 100 | 94.3 |
| MAE | 0.200 | 0 | 0.381 |
| Mean signed diff (bias) | 0.002 | 0 | 0.005 |

### Performance by extraction type

Performance differed substantially by extraction type. In the held-out test set, there were 190 and 210 explicit and inferred cases, respectively. Explicit extraction showed near-perfect raw agreement (ICC = 0.99, n=190, 92.6% exact match). All 14 discordant explicit cases were reviewed against the source note and confirmed to be discordances between the registry and clinical notes, rather than LLM extraction errors, yielding 100% exact agreement (ICC = 1.00) once corrected. Inferred extraction showed lower uncorrected agreement (ICC = 0.72, n=210, 50.5% exact match), rising to ICC = 0.90 (95% CI 0.87 – 0.92) with the adjudicated corrections applied. One confirmed LLM error identified a specific, actionable limitation of the current prompt: a chart in which fever was present but attributable to a concurrent viral illness rather than active JIA.

Among inferred extractions, near-remission cases accounted for a substantial share of discordances. 38 of 104 inferred discordances (36.5%) differed by exactly 0.5 points, and the single most common pattern (n=20) was a registry value of 0 against an inferred value of 0.5.

### Zero-inflation sensitivity analyses

PhGA is zero-inflated in this population, with 50.2% (n=201) charts having a PhGA score of 0 (Supplementary Figure S1). Uncorrected ICC(2,1) restricted to the active disease subset (i.e. PhGA > 0, n=199) was 0.91, higher than the corresponding unrestricted value (ICC 0.87). Because this analysis removes the zero-inflated portion of the distribution entirely rather than reweighting around it, it provides direct evidence that residual disagreement concentrates at the inactive/near-zero boundary rather than reflecting a general difficulty grading disease severity. This is corroborated by treating PhGA=0 versus PhGA>0 as a binary classification: with inactive disease (PhGA=0) as the positive class, sensitivity was 84.1% and specificity was 95.5%, and binary agreement was 89.8% (Cohen’s κ = 0.80).

### Extraction confidence

The LLM’s self-reported confidence field was strongly associated with actual accuracy (Table 4). High- confidence extractions (n=292) achieved ICC(2,1) = 0.93 (exact agreement=86.0%; MAE = 0.200) against registry values, compared with ICC(2,1) = 0.68 (28.7% exact agreement; MAE = 0.801) for medium-confidence extractions (n=108). No cases were rated as low-confidence.

**Table 4.** Agreement stratified by the LLM’s self-reported extraction confidence, using the CARRA Registry as reference standard.

|  | **High** | **Medium** |
|---|---|---|
| **Development set\*** |  |  |
| N | 72 | 27 |
| ICC(2,1) (95% CI) | 0.92 (0.88 – 0.95) | 0.80 (0.62 – 0.91) |
| % Matching gold standard |  |  |
| Exact match | 76.4 | 18.5 |
| Within 0.5 | 80.6 | 59.3 |
| Within 1.0 | 95.8 | 85.2 |
| MAE | 0.292 | 0.722 |
| Mean signed diff (bias) | -0.208 | 0.130 |
| **Test set** |  |  |
| N | 292 | 108 |
| ICC(2,1) (95% CI) | 0.93 (0.91 – 0.94) | 0.68 (0.55 – 0.76) |
| % Matching gold standard |  |  |
| Exact match | 86.0 | 28.7 |
| Within 0.5 | 90.1 | 59.3 |
| Within 1.0 | 94.5 | 82.4 |
| MAE | 0.200 | 0.801 |
| Mean signed diff (bias) | -0.022 | 0.014 |

**NB:** No test-set extractions were assigned low confidence; one development-set extraction was rated low confidence and is not shown.

### Provider-level agreement

Charts were linked to one of 21 distinct providers. Of these, 13 had at least 10 notes in the test set, sufficient for a reasonably stable provider-level ICC estimate; the remaining providers (n=18 charts total) are reported descriptively only, given inadequate precision for individual ICC estimation at that sample size.

Among the 13 providers with n≥10, ICC against the registry-documented gold standard ranged from 0.79 to 0.98 (Supplementary Table S2). No consistent directional bias was observed across providers (mean signed difference ranged −0.14 to +0.28), and individual providers’ largest discordances were distributed in both directions rather than reflecting a uniform pattern, arguing against a single systematic scoring bias tied to provider.

The lowest-agreement provider (ICC = 0.79, n=47) was examined directly via chart review. Notes from this provider were subjectively judged to be more frequently ambiguous in their description of joint findings than notes from higher-agreement providers, offering a plausible explanation for this provider’s lower agreement.

### Multi-run aggregation

Aggregating across runs had a negligible effect on agreement. Using the pre-specified median-of-three rule, ICC rose to 0.88 against the registry, with a median MAE of 0.350.

### Self-consistency Assessment

Single-run repeatability was ICC(2,1) = 0.97 (95% CI 0.97-0.98) across the three runs (pairwise range 0.97–0.98), indicating that a single execution reproduces closely. Extractions were identical across all three runs for 309 (77.2%) charts; the aggregation rule therefore affects only the remaining 91. Median- of-three aggregation changed the extracted value relative to a single run on 7.8–10.2% of charts depending on the run (Supplementary Table S3).

Instability was almost entirely confined to the inferred extractions. Explicit extractions were identical across all three runs in 98.9% of cases (self-agreement ICC = 1.00), compared with 57.6% for inferred extractions (ICC = 0.94). Extraction method itself was unstable in a few cases: 7 charts (1.8%) were assigned a different extraction type in at least one of the three runs. Manual review of these charts found 3 causes. In 4 charts, the note documented a PhGA using a label in the prompt as high-confidence (“MD global”, “Physician VAS”), yet the value was not recognized in at least one run. In one chart, the documented value used a malformed denominator (“1/8”). The remaining two involved ambiguous text: a value labeled only “MD” without indicating a global assessment, which 2 runs treated as explicit and one did not, and the use of non-standard acronym “GPA”.

Cross-run instability was strongly associated with error. Using median-of-three values, charts stable across all three runs had an MAE of 0.233 and exact-match agreement of 81.9%, compared with 0.747 and 31.9% for charts with any between-run disagreement. Instability was likewise associated with model- reported confidence: 90.8% of high-confidence extractions were stable across all three runs versus 40.7% of medium-confidence extractions. Extraction confidence and cross-run instability therefore provide two complementary signals identifying extractions most likely to require manual review, both obtainable without reference to a gold standard. This association was not uniform: the single most unstable chart in the test set (values of 6.0, 2.0, and 5.0 across three runs against a registry value of 5.0) was assigned high confidence, indicating that confidence alone does not fully capture extraction reliability.

### Assessment of hallucinations

Automated screening of all 400 test-set extractions identified no fabricated citations. Blinded manual review of 50 extractions (20 Explicit, 30 Inferred, randomly selected within each stratum) identified no instances of citation, schema, or clinical fabrication, giving an observed hallucination rate of 0% (one- sided upper 95% bound 6.0% by the rule of three; 15.0% within the Explicit stratum and 10.0% within the Inferred stratum).

One reviewed extraction, while meeting all three pre-specified criteria, illustrated a distinct failure mode. The source note documented a swollen and tender second DIP joint but described the patient’s remaining pain regions as of uncertain attribution, offered an alternative explanation for them (prior Osgood- Schlatter disease at the same site), and raised then withdrew the possibility of enthesitis. The extraction’s rationale cited the documented findings and treatment escalation accurately without fabricating any content, but did not account for the clinician’s stated uncertainty. The resulting score exceeded the reference value by one point.

## DISCUSSION

This study demonstrates that an LLM can reliably recover PhGA from routine clinical notes, achieving agreement with an adjudicated reference standard that exceeds previously published inter-rater reliability for the measure.^7^ To our knowledge, this is the first study to show that an LLM can not only extract explicitly documented PhGA values, but also in infer PhGA from clinical narratives when no score is recorded. The capability offers a scalable approach to recover missing PhGA data in clinical registries, expanding the availability of these measures for downstream clinical research.

Beyond recovering missing values, our study found that registry-recorded PhGA differed from the value documented in clinical notes in 14 of 190 explicit extractions (7.4%), pointing to possible errors in existing registry data. The differences ranged from −2 to +2 points. Discrepancies of this magnitude can be consequential: PhGA is a direct component of the clinical Juvenile Arthritis Disease Activity Score (cJADAS10), and based on the American College of Rheumatology 2021 cJADAS10 cutoffs, a 2-point difference can move a visit between disease activity states (e.g. across the inactive disease threshold of 2.5 in polyarticular JIA).^8^ In our analysis, we treated the note as the gold standard where PhGA was explicitly documented, on the premise that it is a more direct record of the physician’s assessment than the registry entry transcribed separately, typically by a research coordinator. However, it is possible that some discordances may reflect documentation error in the note, rather than transcription error in the registry. Reconciling these discrepances would be important beyond the goal of this study.

Among inferred extractions, near-remission cases accounted for a substantial share of discordances. This is likely partially attributable to the prompt design. During development of the prompt, we observed systematic under-scoring of near-remission cases, in which notes documenting remission alongside minor residual findings were assigned a score of 0. We therefore introduced rules assigning 0.5 where remission or inactive-disease language is accompanied by a single minor residual finding, and 1.0 where two or more are present. The 0-versus-0.5 discordances observed in the test set correspond closely to this presentation. Although these rules corrected an error pattern observed in the development dataset, they may have over-corrected in the test set. These rules may also not transfer at another institution, where near-remission findings may be documented differently. Testing in an external cohort with different physicians and documentation practices is needed to determine whether the half-point rules improve accuracy.

Given the well-documented concerns about hallucination in medical applications of LLMs,^9,10^ the absence of detected fabrication in our study is reassuring. This finding may in part reflect the design of the extraction task. Hallucination is most often characterized in the context of open-ended generation, where an LLM generates assertions with no source against which they can be verified.^11^ PhGA extraction is a tightly bounded task: the complete source document (i.e. clinical note) is provided in the context window, the output is short and constrained by a pre-specified schema, and the scoring rules are defined by a closed set of tables, ranges, and preconditions. Importantly, the output schema requires verbatim supporting text before a finding can be asserted, thus forcing the model to identify evidence in the source document rather than generate a claim and justify it afterward. These design features may reduce opportunities for unsupported content generation and are consistent with approaches shown to improve LLM reliability in other settings.^12,13^

Several specific improvements to the prompt are indicated for the next iteration. First, the scoring rules currently do not account for symptoms that may not be attributable to JIA. In one chart, this produced a high-severity score for fever caused by a concurrent viral illness. Second, in the explicit extractions, several labels (e.g. a bare “MD” preceding a fraction, and the acronym “GPA”) were treated inconsistently across runs. These cases should be classified as ambiguous, routing to inference. Another issue relates to how documented uncertainty is handled. Our prompt specified how to weight documented findings but did not specify how to treat findings that the clinician had expressed uncertainty. Specifying this directly (e.g. instructing that findings the clinician does not attribute to JIA with reasonable confidence be excluded from scoring) may be a reasonable refinement for subsequent iterations.

### Limitations and future directions

There are several limitations to our study. First, prompt behaviour may be model-dependent. We optimized our prompt template against a single model version (GPT 5.2). Its performance cannot be assumed to generalize to other models or on a subsequent version of the same model. Second, we conducted the evaluation at a single institution. Some prompt components were developed in response to patterns observed in our development data and may encode local documentation conventions, which may not generalize to other institutions. External validation is therefore an important next step. Some degree of site-specific adaptation, however, may be unavoidable, given differences in documentation conventions across institutions. However, we anticipate that the underlying scoring logic will require less adaptation, given that it is grounded in how JIA is assessed, rather than in the specific ways they are documented.

Finally, clinician-adjudicated scores can only be derived from the content of the note, which may not be complete.

Our analysis points to several specific improvements to prioritize in the next prompt iteration. First, the scoring rules currently do not account for clinical manifestations that resemble JIA-related symptoms but not attributable to underlying JIA disease activity. In one case, the model incorrectly assigned a higher PhGA based on fever caused by a concurrent viral illness rather than JIA disease activity. Second, several non-conventional labels for PhGA (e.g. “MD” preceding a fraction, or the acronym “GPA”) were treated inconsistently across runs. Such cases should be classified as ambiguous and routed to the inference pathway, rather than treated as definitive extractions. Another area for refinement is the handling of documented clinical uncertainty. Although the prompt specified how to weight documented findings, it did not explicitly address findings for which the clinician expressed uncertainty about their attribution to JIA. This could be addressed in subsequent iterations by specifying that findings not attributed to JIA by the clinician with reasonable confidence should be excluded from PhGA scoring.

## CONCLUSION

Our findings demonstrate the potential of LLMs as a scalable approach to addressing gaps in the availability of PhGA for clinical research. With careful design and validation, these approaches could make clinical registries more complete and support their use in downstream research. s

## Data Availability

Electronic health record (EHR) data used in this study are not publicly available due to institutional and patient privacy and confidentiality restrictions. Data from the Childhood Arthritis and Rheumatology Research Alliance (CARRA) Registry may be available upon reasonable request and subject to review and approval by CARRA in accordance with its data access and sharing policies.

## Supplementary Materials

**Supplementary Figure S1.**
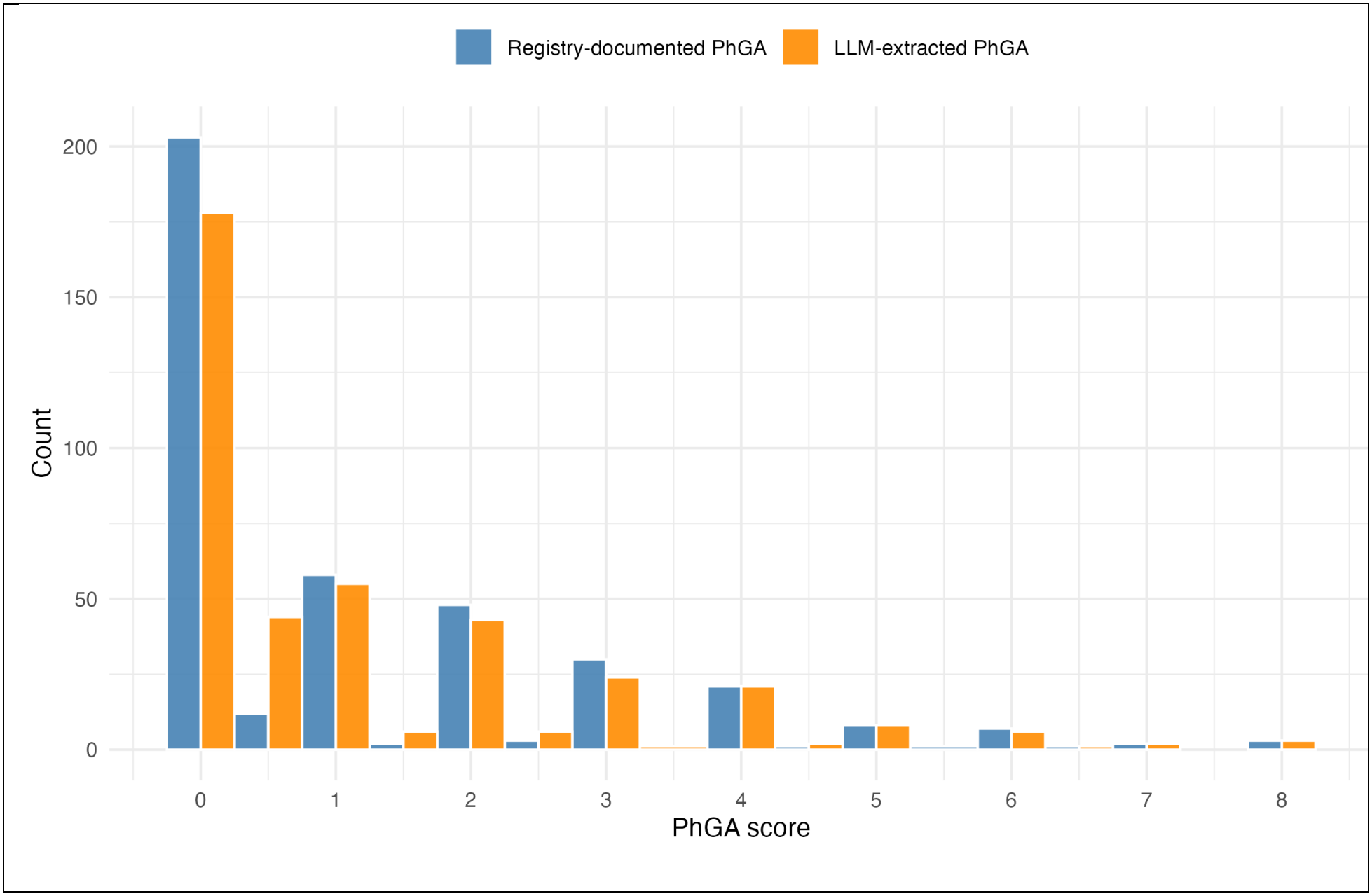
Distribution of PhGA scores in the test set (n=400)

**Supplementary Table S1.** Agreement between LLM-extracted and registry-documented PhGA, stratified by treating provider.

| **Provider** | **Test set** |  |  |  | **Development set** |  |  |  |
|---|---|---|---|---|---|---|---|---|
|  | **N** | **ICC** | **MAE** | **Mean signed diff** | **N** | **ICC** | **MAE** | **Mean signed diff** |
| 4 | 47 | 0.79 | 0.415 | 0.032 | 9 | 0.94 | 0.500 | 0.167 |
| 7 | 16 | 0.86 | 0.500 | 0.188 | 12 | 0.94 | 0.417 | -0.250 |
| 10 | 21 | 0.87 | 0.452 | -0.071 | 5 | 0.92 | 0.600 | 0.200 |
| 17 | 23 | 0.88 | 0.283 | 0.196 | 6 | 0.69 | 0.667 | 0.167 |
| 19 | 12 | 0.89 | 0.208 | 0.208 | 8 | 0.97 | 0.125 | -0.125 |
| 13 | 10 | 0.91 | 0.400 | 0.300 | 4 | 0.95 | 0.375 | -0.125 |
| 15 | 19 | 0.92 | 0.421 | 0.000 | 2 | NA* | NA* | NA* |
| 3 | 4 | 0.92 | 0.250 | 0.250 | 1 | NA* | NA* | NA* |
| 11 | 62 | 0.93 | 0.250 | 0.089 | 13 | 0.87 | 0.269 | -0.269 |
| 16 | 41 | 0.94 | 0.354 | -0.061 | 6 | 0.87 | 1.083 | -0.917 |
| 14 | 30 | 0.95 | 0.283 | -0.217 | 5 | 0.92 | 0.300 | -0.300 |
| 1 | 53 | 0.97 | 0.151 | 0.038 | 14 | 0.95 | 0.321 | -0.036 |
| 2 | 20 | 0.98 | 0.050 | 0.000 | 7 | 0.95 | 0.143 | 0.143 |
| 9 | 28 | 0.98 | 0.125 | 0.054 | 6 | 0.96 | 0.333 | -0.333 |
| 18 | 5 | 1.00 | 0.000 | 0.000 | 2 | NA* | NA* | NA* |
| 23 | 2 | NA* | NA* | NA* | 0 | NA* | NA* | NA* |
| 5 | 2 | NA* | NA* | NA* | 0 | NA* | NA* | NA* |
| 22 | 0 | NA* | NA* | NA* | 0 | NA* | NA* | NA* |
| 20 | 2 | NA* | NA* | NA* | 0 | NA* | NA* | NA* |
| 21 | 2 | NA* | NA* | NA* | 0 | NA* | NA* | NA* |
| 6 | 1 | NA* | NA* | NA* | 0 | NA* | NA* | NA* |

\* Providers with fewer than 3 comparisons are excluded from ICC estimation given inadequate precision at that sample size and are marked NA

**Supplementary Table S2.** Multiple runs and aggregation comparison.

|  | **ICC** | **Exact match (%)** | **MAE** | **Bias** |
|---|---|---|---|---|
| **Run 1** | 0.87 | 70.5 | 0.363 | -0.0125 |
| **Run 2** | 0.86 | 70.2 | 0.380 | -0.0375 |
| **Run 3** | 0.87 | 69.5 | 0.366 | -0.0219 |
| **Median** | 0.88 | 70.5 | 0.350 | -0.0250 |

